# COST AND UTILIZATION TRENDS OF ENDOMYOCARDIAL BIOPSY IN HEART TRANSPLANT PATIENTS: A 4-YEAR CLAIMS DATA ANALYSIS

**DOI:** 10.1101/2023.09.13.23295491

**Authors:** Adrian Vilalta

## Abstract

**Objective:** This study evaluated patterns of utilization, complications, and costs of endomyocardial biopsies (EMB) in heart transplant patients.

**Methods:** The IBM® *Treatment Pathways®* tool was used to analyze claims data selected from the IBM®’s *MarketScan®* de-identified, HIPAA-compliant dataset. Differences in EMB paid amounts and utilization patterns were assessed for commercial payers and Medicare for years 2016 to 2019. Type, frequency, and overall cost of complications of the EMB procedure in these patients were also evaluated.

**Results:** A total of 8,170 records (6,385 commercial payers and 1,785 Medicare) of heart transplant patients with evidence of EMB procedures performed between 2016 and 2019 were identified in the database. In 2019, the median paid amount for an outpatient EMB in a heart transplant patient was US $7,918 (commercial) and US $2,980 (Medicare). Heart transplant patients received between 4.6 and 6.8 (median; Medicare, commercial) EMBs the first year after the transplant. Approximately 25% of EMB procedures were associated with complications. In 2019 the total cost of EMB complications per patient was US $9,049.

**Discussion:** Analysis showed that the paid amount for the EMB procedure increased by almost 25% from 2016 to 2019 for commercial payers. Given the high frequency of complications after the EMB procedure and the associated cost of the complications it is estimated that the median paid amounts are closer to US $10,000 per patient per EMB. Given the number of EMBs provided, the associated risks, and the paid amount trends, non-invasive alternatives to EMB should be considered for the surveillance of heart transplant patients.

## INTRODUCTION

Heart transplantation is a lifesaving, last resort measure for people with severe heart disease. According to the Organ Procurement & Transplantation Network (OPTN), there were 3,552 heart and 45 heart-lung transplants in 2019 in the United States.^1^ Approximately 37,000 patients are alive today with a heart transplant.^2^

The prognosis for heart transplant patients has greatly improved since the first heart transplantations in the late 1960s. Nowadays, approximately 85 to 90% of heart transplant patients are living one year after their surgery; the five-year survival approaches 80%.^2^ On the other hand, heart transplantation is a specialized, costly procedure. According to a recently published analysis, a heart transplant incurs approximately US $1,664,800 in billed charges.^3^ In addition, complications from the organ transplant are not uncommon and can generate up to an additional US $250,000 in yearly charges.^4^ Therefore, maintaining the heart transplant patient’s health and maximizing the useful life of the graft requires close and careful monitoring.

Endomyocardial biopsy (EMB) has been routinely used to monitor cardiac allograft rejection. The frequency of surveillance EMBs is typically greatest during the first three to six months after transplantation, the time at which acute cellular rejection (ACR) is most common.^4^ Recent data from a multi-center trial suggest that heart transplant patients typically get 5 to 6 EMBs the first year after the transplant.^5^ However, there is variability in the number of EMBs that patients receive, with some organ recipients receiving over 20 biopsies in a year. This is a well-recognized pain point in the heart transplant patient’s journey. Recent (Jan 2021) comments by Dr. Sean Agbor-Enoh, a leading researcher from the National Heart, Lung, and Blood Institute (NHLBI) highlight this fact. Dr. Agbor-Enoh stated: “there is an urgent need for an alternative method to monitor patients for acute heart transplant patients”.^6^

Heart biopsies are invasive and painful. Their routine use can also lead to the occlusion of access points and damage to the allograft. In addition, the EMB event incurs ancillary medical costs such as patient sedation and monitoring, among others. Moreover, published studies have shown that biopsies have several severe limitations including sampling error and inter-pathologist variability which make EMB a flawed standard of care.^5^

Biopsies require a visit to a specialized transplant center which may expose the immuno-suppressed patient to opportunistic infections including SARS-CoV-2/COVID-19.^7^ Given this risk, the International Society for Heart and Lung Transplantation (ISHLT) has issued recommendations to minimize the risk of infection while properly maintaining cardiac graft health surveillance. The ISHLT recommends the use of non-invasive and telehealth options whenever possible.^8^ This guidance is aligned with recommendations from the Centers for Disease Control and Prevention (CDC).^7^

Despite being used routinely, there is a lack of current, real-world data on the financial cost of EMBs, especially as it pertains to the heart transplant population. Existing published cost effectiveness analyses use cost data listed in the Centers for Medicare & Medicaid Services (CMS) fee schedule which does not reflect the reality of the paid amounts by commercial insurers.^9,10^ Importantly, actual payment data are required as billed charges overestimate actual payment for services. Real-world data on the costs and risks of EMBs is an essential input for the development of meaningful cost-effectiveness analyses of EMB and alternative surveillance modalities.

The present study aimed to generate accurate, current, and complete data on the financial cost in US dollars of an outpatient endomyocardial biopsy event by payer type. Additionally, real-world data on the frequency and economic impact of complications from EMBs were also developed.

## MATERIALS AND METHODS

Data presented here were obtained through a retrospective, observational claims-based analysis using a de-identified, HIPAA-compliant database to describe the patterns of utilization and episode costs of endomyocardial biopsies in heart transplant patients.

### Record Selection

Insurance claims from heart transplant patients with evidence of an EMB procedure and enrollment in either a commercial or Medicare insurance plan were identified through the IBM® *MarketScan*® *Treatment Pathways*® database and analytical interface.^11^ For additional context, claims data for EMB procedures with no evidence of a heart transplant, as well as claims data for heart transplant patients without selection for EMB were also obtained. *MarketScan*® collects all the coordination of benefits (COB) data thus generating total amount paid information, ie, insurance, co-insurance, and patient pay. The healthcare event comprised all procedures performed the day of the EMB. The cohort was further refined by selecting records with evidence of heart transplantation. We obtained data for calendar years 2016 through 2019. During that period, the IBM® *MarketScan*® database contained the de-identified annual pooled healthcare data of close to 200 million individuals insured commercially, or as part of the national distributed Supplemental Medicare and Medicaid programs throughout the entire United States. Using the approach described above allows for the generation of fully de-identified, HIPAA-compliant data; therefore, no IRB review was required.

### Study Measures

Evidence of EMB was established using Current Procedural Terminology (CPT) codes. The CPT code set is maintained by the American Medical Association.^12^ Diagnosis of heart transplantation was established using International Classification of Diseases (ICD-10) codes. ICD codes are created by the World Health Organization and contain codes for diseases, signs and symptoms, abnormal findings, complaints, social circumstances, and external causes of injury or diseases.^13^ Details on codes used in the present study can be found in **SUPPLEMENT 1**.

### Limitations

As with other claims-based studies, the findings may be impacted by the sensitivity of the claims data. Cohorts defined through this analysis do not consist of the complete universe of patients receiving the procedure. Only outpatient claims were included in the present analysis therefore costs of episodes systematically omit any inpatient services incurred through treatment of complications. Claims data may underestimate health care utilization among low-income individuals including Medicaid-insured and the non-insured population.

## RESULTS AND DISCUSSION

The number of records of heart transplant patients with evidence of EMB identified by our analysis are presented in **Figure 1**. According to the Organ Procurement & Transplantation Network (OPTN), there were 3,191 heart transplants in 2016 and 3,552 in 2019.^1^ Our count of heart transplant patients with evidence of an EMB range from 1,798 (56% of OPTN) in 2016 to 2,229 in 2019 (62% of OPTN) clearly representing a significant proportion of the total number of heart transplants reported every year.

**Figure 1.**
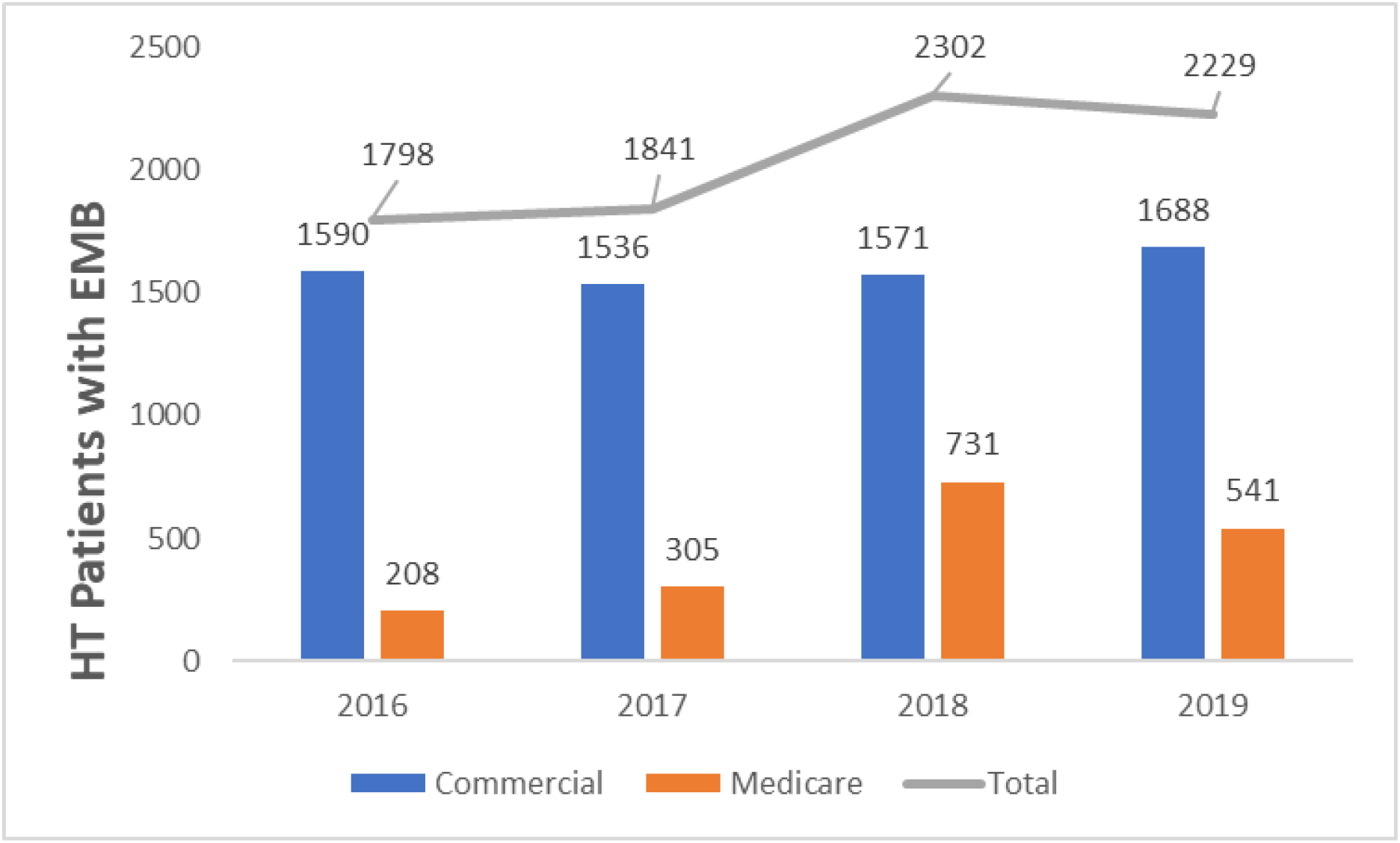
Endomyocardial biopsy records 2016 to 2019. Number of heart transplant patient claims with evidence of endomyocardial biopsy (CPT 93505 and evidence of heart transplant) broken down by payer type. Total number of claims identified for each year is shown above the trendline.

### Number of EMB received

Our analysis also revealed that heart transplant patients received 4.6 to 6.8 EMBs (Medicare, commercial) during the first year after the transplant. The frequency of EMBs established by our analysis is in line with published estimates. The frequency of surveillance EMBs is typically greatest during the first three to six months after transplantation, the time at which acute cellular rejection (ACR) is most common.^5^ Recent data from a multi-center trial suggest that heart transplant patients typically get 5 to 6 EMBs the first year after the transplant.^5^

### Paid amount for EMB

Paid amounts per EMB episode are summarized in **Table 1**. Our analysis indicates that the amount paid for an EMB by commercial payers has increased steadily with a median of US $6,341 paid in 2016 to US $7,918 paid in 2019, representing close to 25% increase over four years. On the other hand, Medicare paid close to 30% less for an EMB in 2019 compared to 2016. Our findings on the paid amounts for EMB procedures by Medicare are consistent with those published by others.^9,10^ However, our literature searches did not identify any recent publications where the paid amounts for the EMB procedure by commercial insurers was reported. We believe that the claims data for commercial plans should be considered in future cost-effectiveness analyses given the proportion of procedures covered by commercial insurance and the significant difference in the amount paid as compared to Medicare.

**TABLE 1.**

| Endomyocardial Biopsy -<br>Paid Amounts per Episode of Care (US \$) | | |
| --- | --- | --- |
|  | Mean | Median |
| **2016** |  |  |
| Commercial Insurance | \$8,357 | \$6,341 |
| Medicare | \$5,214 | \$3,908 |
| **2017** |  |  |
| Commercial Insurance | \$8,056 | \$6,647 |
| Medicare | \$7,355 | \$3,302 |
| **2018** |  |  |
| Commercial Insurance | \$9,285 | \$6,731 |
| Medicare | \$5,350 | \$3,375 |
| **2019** |  |  |
| Commercial Insurance | \$9,052 | \$7,918 |
| Medicare | \$8,572 | \$2,980 |

### Services performed with the EMB

We performed a proximate event analysis using the code for endomyocardial biopsy (CPT 93505) as the index code. This analysis identified procedures associated with CPT 93505 and their frequency the day of the EMB procedure. The proximate event analysis was done to better understand the composition of procedures contributing to the total paid amount of the EMB episode of care. The list of the top 25 codes co-appearing in the EMB claims is provided in **Table 2**.

**TABLE 2.**
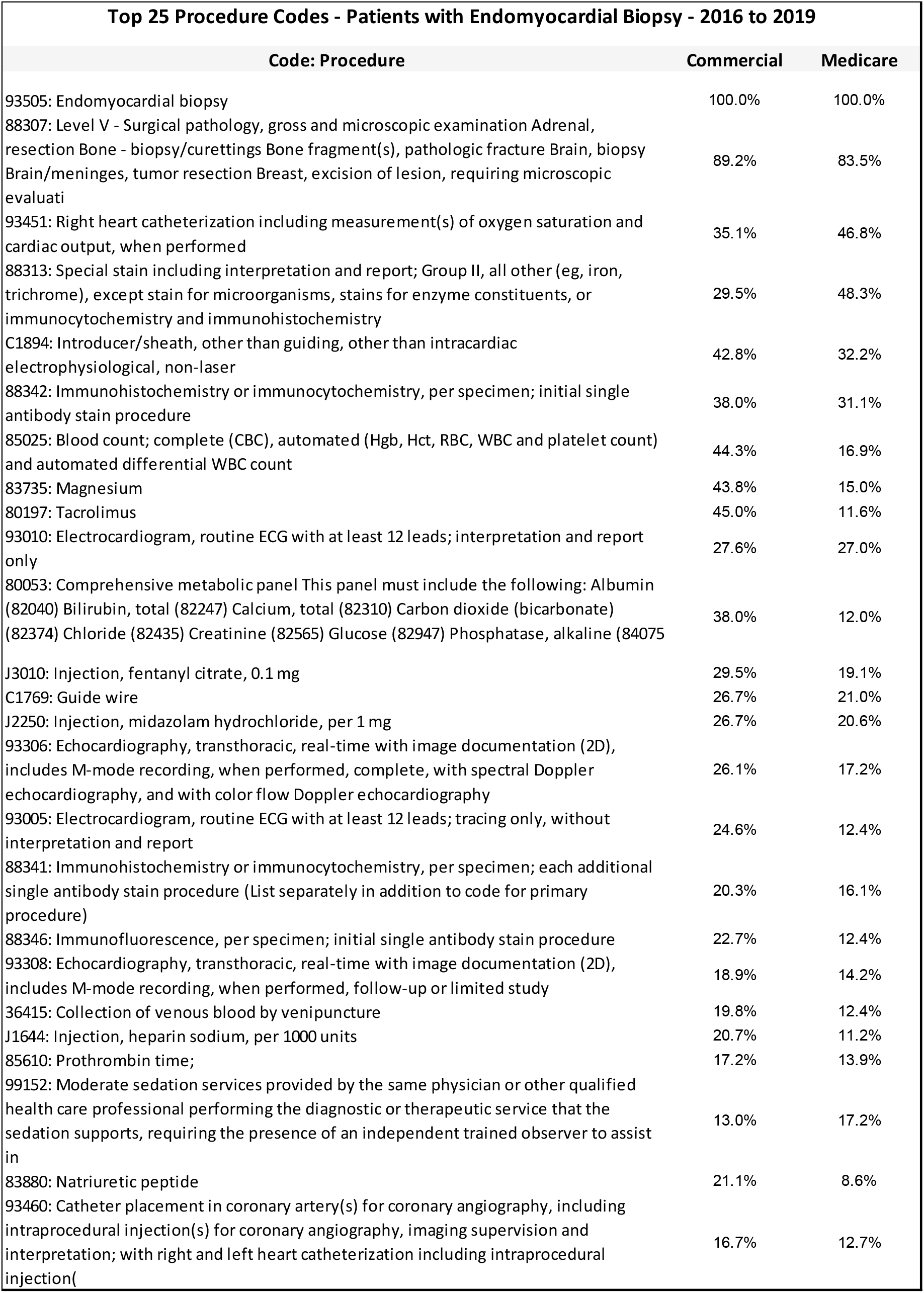
25 MOST FREQUENT CODES CO-APPEARING IN ENDOMYOCARDIAL BIOPSY CLAIMS.

### EMB complications and cost

According to our findings up to 25.2% of patients receiving an EMB will experience post-procedure complications. The most frequent complications are nonrheumatic tricuspid (valve) insufficiency (45.6 to 49.7%), cardiac arrhythmia (23.8 to 26.2%), and atrioventricular block (4.8 to 9.7%). A full list of the most frequent reported complications can be found in **Table 3**. The frequency of complications revealed by our analysis differs from that published by others on the safety of EMB. For example Yilmaz *et al*. report a rate of EMB complications of 2.24% to 5.10% for right ventricular endomyocardial biopsy.^14^ Also, a widely cited article by Cooper *et al*. reports a frequency of EMB complications of under 6%.^15^ However, the rate of complications after EMB reported by these authors corresponds to that observed primarily in patients diagnosed with either myocarditis or cardiomyopathy of unknown origin, not those with a heart transplant. A complication that is relatively unique to cardiac transplant recipients, presumably because of the performance of biopsies in which the bioptome is repeatedly passed across the tricuspid valve, is tricuspid regurgitation (TR). The reported prevalence of TR ranges from 47 to 98 percent and is moderate to severe in as many as one-third of patients.^16,17^ Our findings agree with these studies, highlighting the higher risk of complications of EMB in heart transplant patients. Our analysis also revealed that the median total annual cost of complications of an EMB reached US $9,049 in 2019. This cost should be factored in the median paid amount for an EMB given that approximately one in four heart transplant patients that receive an EMB will experience complications from the procedure. With this in mind, in our estimate, the true cost of an EMB to commercial payers is close to US $10,000 and US $3,771 to Medicare.

**TABLE 3.** FREQUENCY OF COMPLICATION CODES – ALL PATIENTS WITH A HEART TRANSPLANT AND AN ENDOMYOCARDIAL BIOPSY.

| Frequency of Complication Codes - Patients with Endomyocardial Biopsy and Heart Transplant |  |  |
| --- | --- | --- |
| Code: Complication | Commercial | Medicare |
| Patients with diagnosis for complication of endocardial biopsy post biopsy (N, %) |  |  |
| I361: Nonrheumatic tricuspid (valve) insufficiency | 49.7% | 45.6% |
| I499: Cardiac arrhythmia, unspecified | 23.8% | 26.2% |
| I442: Atrioventricular block, complete | 4.8% | 9.7% |
| I2699: Other pulmonary embolism without acute cor pulmonale | 7.1% | 4.9% |
| I82409: Acute embolism & thrombosis of unspec deep vein of unspec low extremity | 6.4% | 5.8% |
| I770: Arteriovenous fistula, acquired | 2.6% | 1.9% |
| I314: Cardiac tamponade | 1.5% | 1.9% |
| I360: Nonrheumatic tricuspid (valve) stenosis | 1.5% | 2.9% |
| I9789: Oth postprocedural comps & disorders of circulatory system NEC | 1.5% | 0.0% |
| I369: Nonrheumatic tricuspid valve disorder, unspecified | 1.4% | 1.0% |
| J95811: Postprocedural pneumothorax | 1.3% | 1.9% |
| I368: Other nonrheumatic tricuspid valve disorders | 0.6% | 1.9% |
| I9751: Accident punct & laceration circ syst organ/structure during circulat px | 0.4% | 0.0% |
| I2690: Septic pulmonary embolism without acute cor pulmonale | 0.3% | 0.0% |
| I2692: Saddle embolus of pulmonary artery without acute cor pulmonale | 0.3% | 0.0% |
| T82827A: Fibrosis of cardiac prosth devices, implants & grafts, initial encounter | 0.3% | 0.0% |
| I362: Nonrheumatic tricuspid (valve) stenosis with insufficiency | 0.1% | 0.0% |
| I97710: Intraoperative cardiac arrest during cardiac surgery | 0.1% | 0.0% |
| J95812: Postprocedural air leak | 0.1% | 0.0% |
| S2619XD: Other injury of heart without hemopericardium, subsequent encounter | 0.1% | 0.0% |
| S2690XA: Unspec injury heart, unspec w or w/o hemopericardium, initial encounter | 0.1% | 0.0% |

## CONCLUSIONS

Endomyocardial biopsy is a common, recurrent procedure used to monitor the health of heart allografts as well as to help diagnose potential complications. Spite its wide use, there is a paucity of recent, real-world data on paid amounts for the procedure, as well as the total cost of the episode of care. This is particularly relevant as it relates to paid amounts by commercial insurers. Also, there is an incomplete understanding of the frequency and economic impact of the clinical risks of the endomyocardial biopsy procedure in the heart transplant population. These data are essential for the evaluation of cost-effectiveness for emerging, non-invasive clinical interventions to manage heart transplant patients.

The present analysis bridges the data gap by providing paid amounts for the complete episode of care by payer type. The records identified through our analysis represent a sizable proportion of all heart transplants in the US and are therefore expected to accurately reflect the situation across the country. In addition, the study provides details on the associated risks of endomyocardial biopsies as well as their economic impact. The cost and patterns of utilization established here provide a solid foundation for follow up analyses of the economic value of novel cardiac allograft surveillance strategies.

## Data Availability

All data produced in the present study are available upon reasonable request to the authors.

---

**SUPPLEMENT 1.**
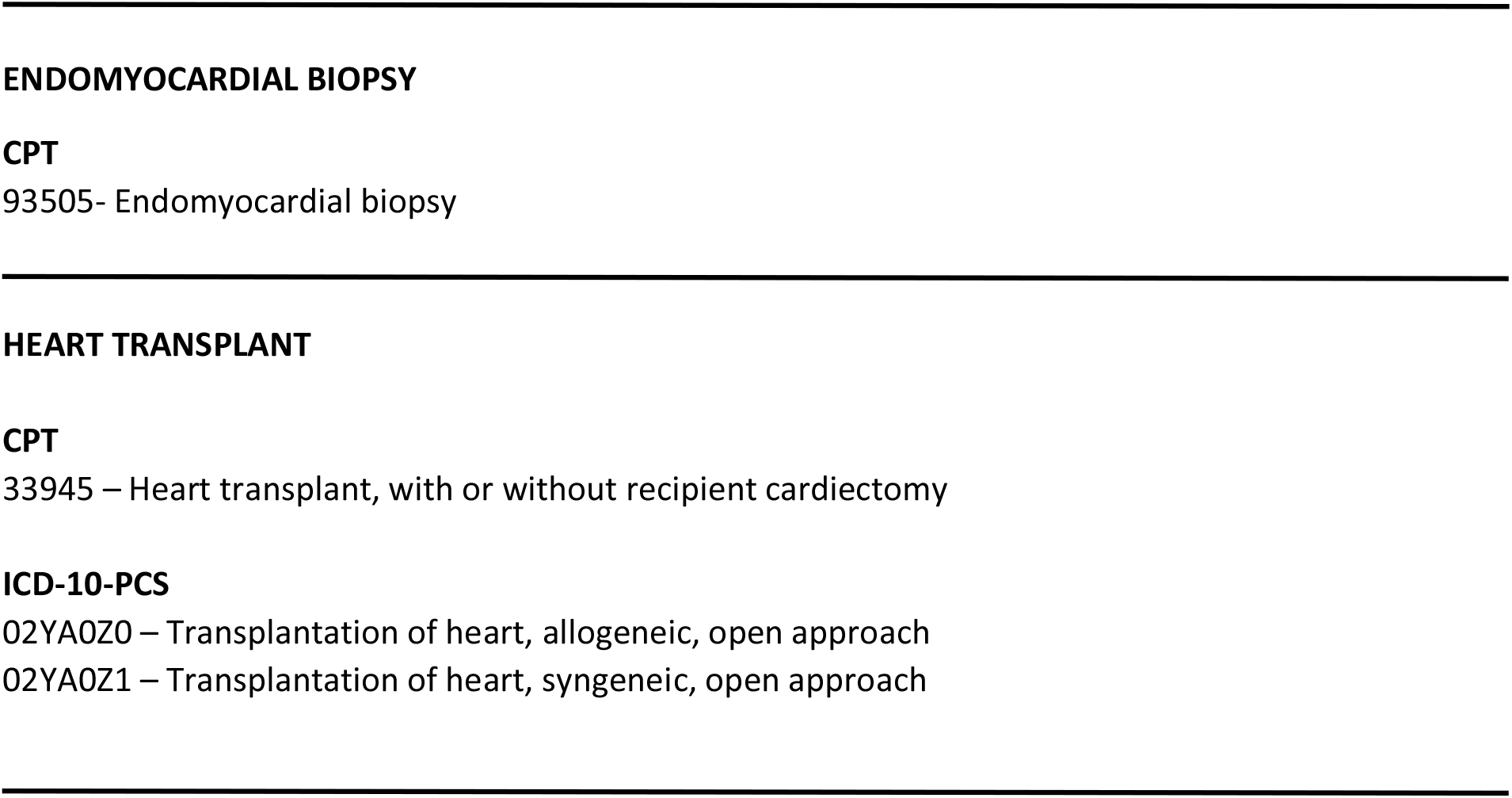
CODES USED TO IDENTIFY CLAIMS DATA.

